# An Outpatient Telehealth Elective for Displaced Clinical Learners during the Coronavirus Pandemic

**DOI:** 10.1101/2020.07.28.20146522

**Authors:** Alec M. Weber, Anoushka Dua, Kitae Chang, Hamsitha Jupalli, Farsha Rizwan, Abhishek Chouthai, Catherine Chen

**Author notes:** Corresponding author: **Alec M. Weber**, Robert Wood Johnson Medical School, Rutgers, The State University of New Jersey, 675 Hoes Lane West, Piscataway, NJ 08854, USA;.

## Abstract

**Introduction:** In response to the Coronavirus pandemic, medical schools suspended clinical rotations. This displacement of students from the wards has limited experiential learning opportunities. Concurrently, outpatient practices are experiencing reduced volumes of in-person visits and shifting towards virtual healthcare. This transition comes with its own logistical challenges. Here, we discuss a workflow that enabled students to engage in meaningful clinical education while helping the RWJMS outpatient practices implement remote telehealth visits.

**Methods:** A four-week virtual elective was designed to offer clerkship students the opportunity to participate in virtual telehealth patient encounters. Students were prepared with EMR training and introduced to an outpatient workflow that supports healthcare providers in the ambulatory setting. Patients were consented to telehealth services before encounters with students. All collected clinical information was documented in the EMR, after which students transitioned patients to a virtual Doxy.me video call appointment. Clinical and educational outcomes of students’ participation were evaluated.

**Results:** Survey results showed students felt well-prepared to initiate patient encounters. They also expressed comfort while engaging with patients virtually during telehealth appointments. Students further identified educational value, citing opportunities to develop patient management plans consistent with in-person clinical experiences. A significant healthcare burden was also alleviated by student involvement. Over 1000 total scheduled appointments were serviced by students who transitioned over 80% of patients into virtual provider waiting rooms.

**Discussion:** After piloting this elective with rising fourth-year students, pre-clerkship students were also recruited to act in a role normally associated with clinical learners (e.g., elicit patient histories, conduct a review of systems, etc.). An additional telehealth elective is being designed so medical students can contribute to inpatient care without risk of exposure to SARS-CoV-2. These efforts are designed to allow students to continue with their clinical education during the pandemic.

## INTRODUCTION

On March 17th, 2020, the AAMC issued strong guidance that medical schools suspend clinical rotations during the Coronavirus pandemic, effective immediately.^1^ A temporary and unified suspension by all institutions would provide the medical education community with time to ensure the safety of their displaced clinical learners as new information about COVID-19 continued to emerge. Medical schools following this guidance au courant are developing innovative curricular modifications to deliver uncompromised education during the ongoing pandemic. Over the past ten weeks, Rutgers Robert Wood Johnson Medical School (RWJMS) has successfully introduced a new telehealth elective designed as a unique educational opportunity to use telehealth technologies for healthcare delivery in service of Robert Wood Johnson Medical Group (RWJMG) patients.

Under normal circumstances, the RWJMG outpatient practices care for approximately 15,000 patients each week. The Commonwealth Fund estimated outpatient practices were scheduling less than half their normal volume of in-person visits throughout March to mitigate community transmission of the novel Coronavirus.^2^ For the numerous practices struggling to care for their patient population, the ability to extend patient-provider relationships beyond traditional in-person visits is invaluable. We developed a telehealth workflow in coordination with RWJMG practice managers to facilitate efficient diagnosis, treatment, management, and education as providers embraced the virtual setting. Students were instrumental during this transition and ultimately reduced a significant workforce burden whilst gaining valuable exposure to a modern healthcare intervention.

## METHODS

### Objectives

Rising fourth-year medical students were recruited throughout March and April 2020 to pilot this elective for a four-week period. To ensure satisfaction of curricular competencies, several objectives were defined for students to: (1) understand the broad applications of telehealth interventions in the outpatient setting, (2) develop fluency with telehealth technologies to support virtual appointments, (3) learn from clinicians and implement techniques to interact with patients during virtual appointments, (4) use telehealth modalities for assessment of clinical status and risk of decompensation, (5) develop a plan of care and counsel patients throughout virtual appointments, and (6) communicate with patients about health literacy and other social determinants of health that may affect the success of virtual appointments. These objectives further served as guidance for the delivery of high-quality patient care in the virtual setting.

### Outpatient Workflow

Students attended mandatory training sessions to become familiar with our electronic medical record (EMR) and the workflow (Figure 1) described as follows. When attending physicians request student support, a Microsoft Excel workbook is populated with their contact information and deidentified appointment times in half-day increments. Automated emails are generated throughout the week to alert students as the Excel workbook populates with new attending physicians and their respective appointment times. Students can indicate availability to support attending physicians via a hyperlink provided in these emails. We also require that students contact each respective attending physician and confirm their request for student support before servicing any patients.

**Figure 1.**
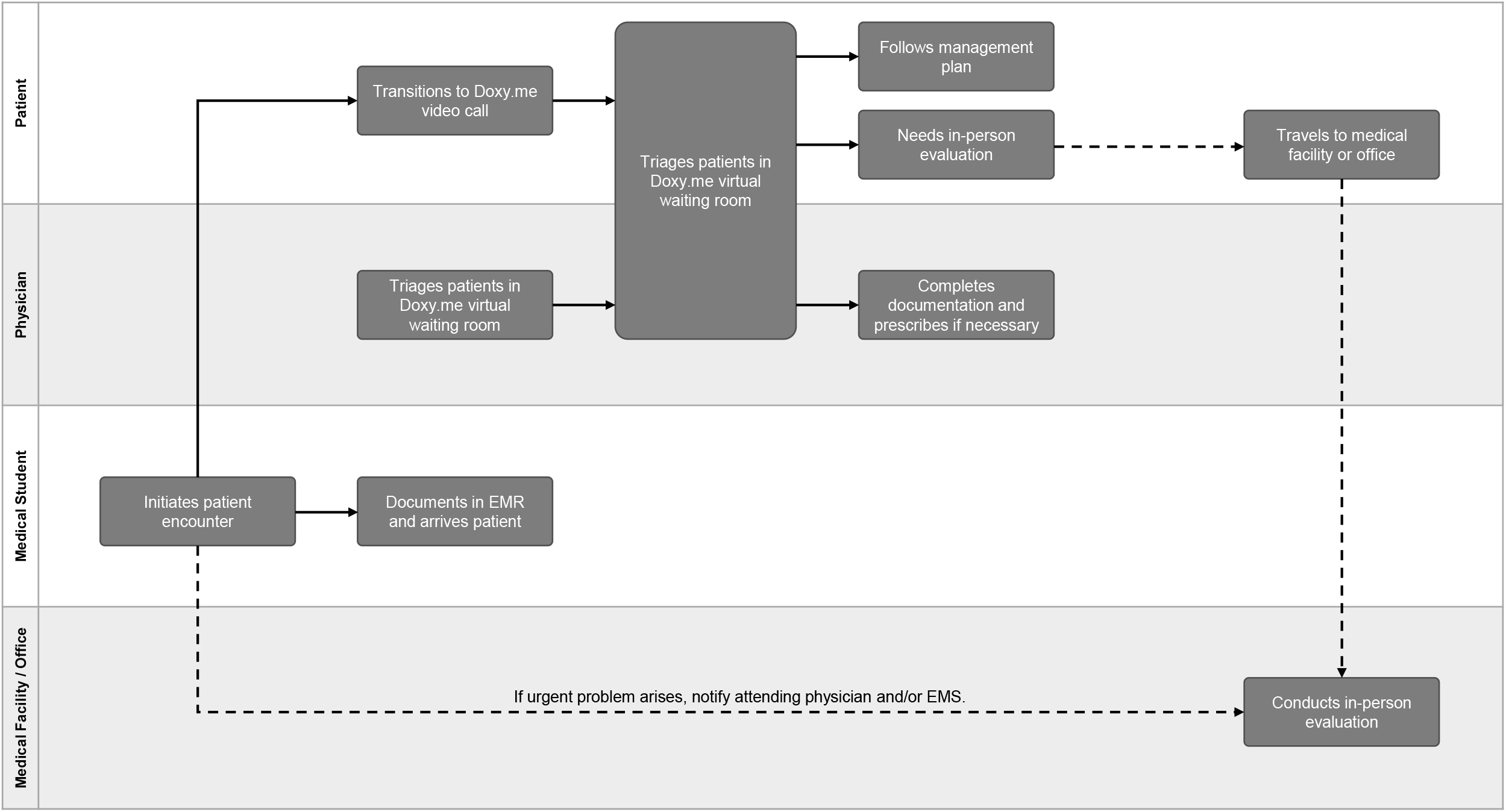
Swimlane diagram of the outpatient workflow developed in coordination with RWJMG practice managers.

Students initiate contact with patients by phone call approximately 30 minutes before their scheduled appointment time. The dialer feature of the Doximity application is used to protect personal phone numbers, and the subsequent patient encounter is documented in the EMR via remote access. A new document type was created in the EMR for telehealth patient encounters to satisfy documentation requirements. This telehealth note requires information about the visit type (phone call or video call), the method used to connect the patient (phone, Doxy.me, FaceTime, other), the patient’s location (state), and the attending physician’s location (clinical office, Rutgers location other than clinical office, home or other non-Rutgers location). Before collecting any clinical information, the patient is provided with information about the risks and benefits of telehealth services. Their verbal consent to participate in telehealth services is obtained and students proceed with the patient encounter thereafter. A chief complaint and history of present illness are collected first, after which a review of systems and medication reconciliation are conducted. Vital signs are then obtained from personal home health monitoring equipment if available (e.g., scales, thermometers, blood pressure monitors, etc.). All clinical information is entered into the telehealth note where applicable.

Next, students transition the patient encounter from the initial phone call to a Doxy.me video call. This HIPAA-compliant telehealth system provides a cloud-based video conferencing platform with a virtual waiting room feature for providers to triage and connect with patients at their appointment time. Students use skills attained from the training sessions to troubleshoot technological difficulties and other issues that may arise during the transition. Students hold their preliminary note in the EMR for use by the attending physician and exit the patient encounter once this transition to Doxy.me is successful. Debrief sessions are held by select attending physicians after certain patient encounters to provide feedback and educate students where appropriate.

To be eligible for elective course credit, students must complete a survey each time they log contributed hours. The survey encompasses barriers to successful telehealth utilization. Various appointment and workflow data are collected to enable retrospective analysis and real-time process improvement. Two weeks of elective course credit are granted per 40 hours of support contributed. Students will write a one-page reflection on their experience with telehealth and virtual patient care.

## RESULTS

### Educational Outcomes

Our previously defined educational objectives were not strictly measured. Rather, this elective was optimized to serve patients of the RWJMG outpatient practices. Surveyed students provided positive informal feedback about the workflow, which enabled confident use of the EMR and telehealth technologies. Many felt they were able to explore the capabilities of virtual patient encounters while receiving practical instruction about various aspects of telehealth. Given the large selection of participating specialty and subspecialty attending physicians at RWJMG, students were able to serve a broad spectrum of patients. Students who supported attending physicians with fewer scheduled patients were able to develop patient management plans and participate extensively during the patient encounter. Students also found interviewing patients, collecting a history, and documenting the encounter all provided significant educational value.

Surveys also proved an invaluable source of information for process improvement. Students were required to enumerate any barriers to initiating patient encounters. They learned how to assist patient with these barriers to ensure a successful telehealth visit. This enumeration of barriers further afforded the opportunity to modify our workflow and better meet the needs of patients.

### Clinical Outcomes

Over a four-week period (April 13 to May 8), 58 attending physicians from 3 specialties (internal medicine, pediatrics, neurology; inclusive of 17 total subspecialties) requested support from 64 rising fourth-year medical students for 1411 total scheduled appointments (Figure 2). In the first week (April 13 to 17), students serviced 173 (80.9%) of 214 total scheduled appointments and transitioned 139 (80.3%) patient encounters to a Doxy.me video call. In the second week (April 20 to 24), students serviced 325 (87.1%) of 373 total scheduled appointments and transitioned 256 (78.8%) patient encounters to a Doxy.me video call. In the third week (April 27 to May 1), students serviced 383 (87.4%) of 438 total scheduled appointments and transitioned 318 (83.0%) patient encounters to a Doxy.me video call. In the fourth week (May 4 to 8), students serviced 337 (87.3%) of 386 total scheduled appointments and transitioned 281 (83.4%) patient encounters to a Doxy.me video call.

**Figure 2.**
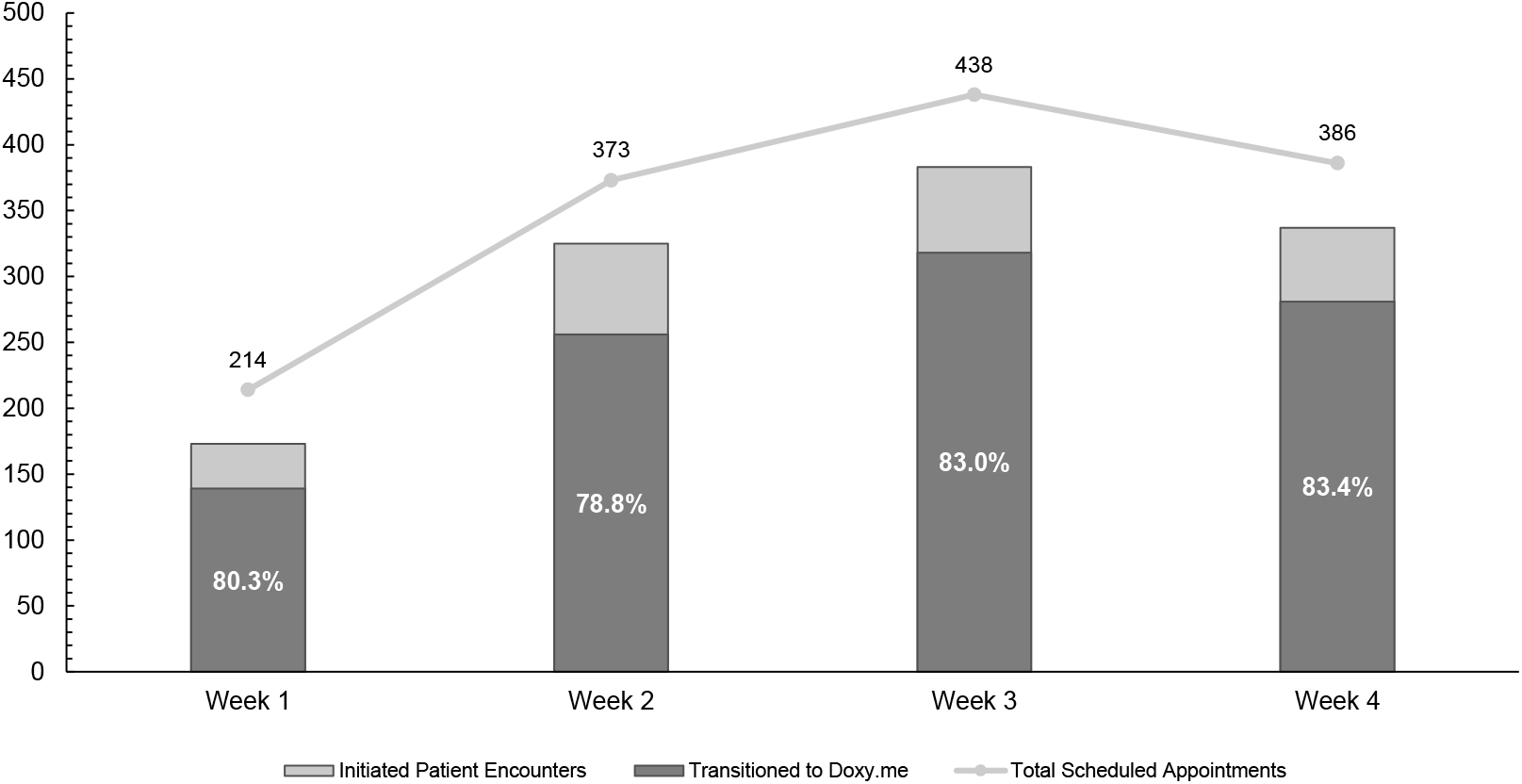
Trend of total scheduled telehealth appointments over a four-week period (April 13 to May 8) for RWJMG specialty clinics (internal medicine, pediatric, and neurology). All patient encounters successfully transitioned to a Doxy.me video call (dark grey) are illustrated as a percentage of initiated patient encounters (light grey).

We encountered several barriers (Figure 3A, B) to successful telehealth utilization during our four-week pilot period. Together, 64 students were unable to initiate the patient encounter for 197 (13.9%) of 1411 total scheduled appointments. We identified two primary reasons: patients either asked to reschedule (80, 41.5%) or simply did not answer the phone (79, 40.9%). Of the 1214 patient encounters that students were able to initiate, 992 (81.7%) were successfully transitioned to a Doxy.me video call. Most patients from the remaining 222 encounters specifically requested transition to a traditional phone call (101, 46.5%) or reported inability to access a compatible smartphone and/or computer at the time (71, 32.7%). Despite assistance from students, 40 (18.4%) patients were not adept enough with technology to enter the Doxy.me virtual waiting room.

**Figure 3.**
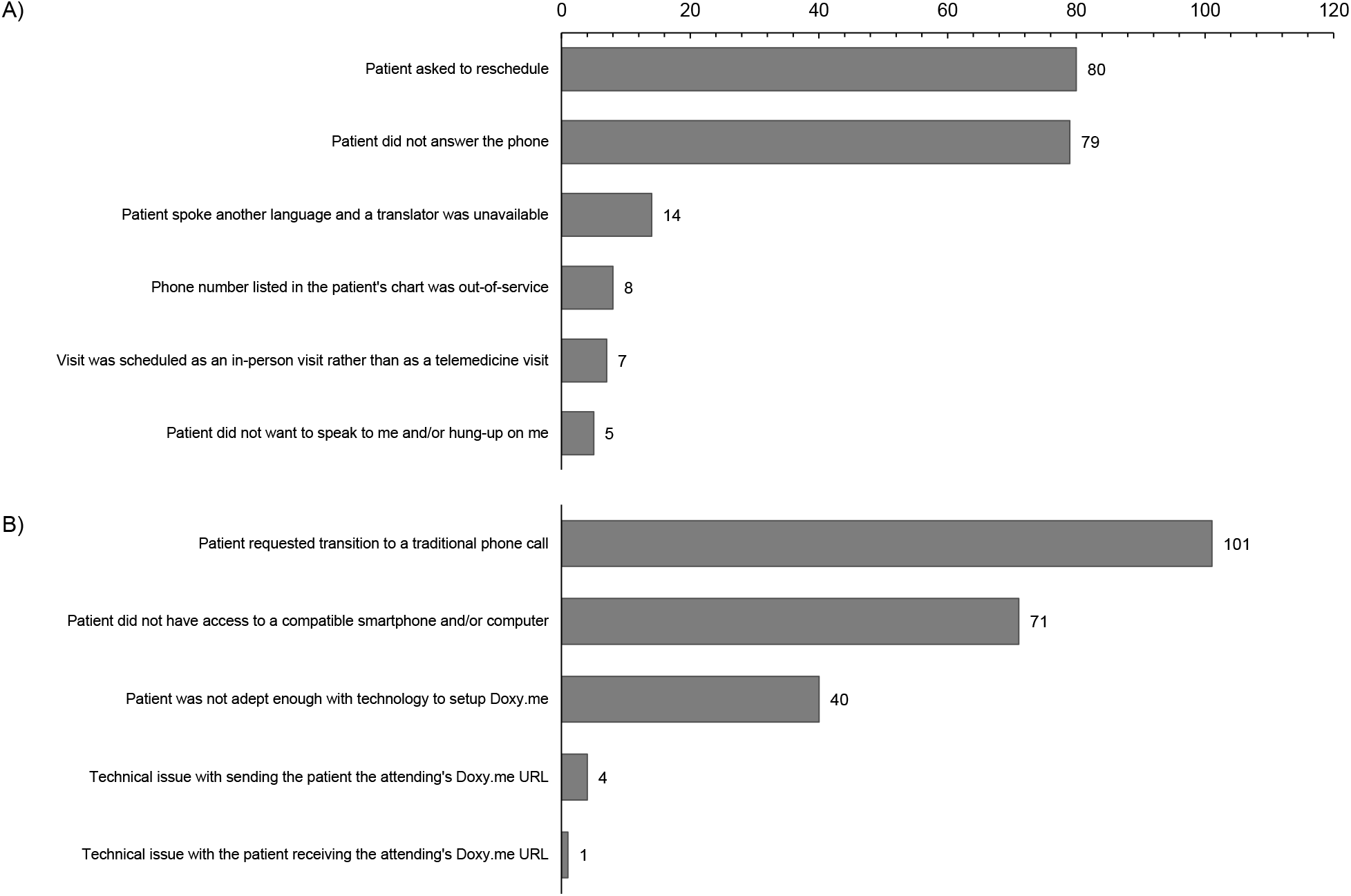
Student survey responses that quantify instances where students were unable to A) initiate the patient encounter and B) transition initiated patient encounters to a Doxy.me video call.

### Limitations

Among the education outcomes, we note several limitations with uniformity. Attending physicians were able to personalize their requests for student support, with some permitting student participation during the actual patient encounters. Although all attending physicians were asked to conduct debrief sessions when possible, time constraints and schedule conflicts reduced the number of educational moments within this telehealth elective.

Among the measured clinical outcomes, data collected about barriers to telehealth implementation are likely skewed. Our pilot was conducted while the RWJMG outpatient practices existed in an unprecedented state of flux without their usual automated scheduling systems in place. Instances where students were unable to initiate the patient encounter via phone call were increasingly corrected towards the end of the pilot as these RWJMG systems were reimplemented. Unfortunately, we were unable to follow up with these patients who were rescheduled to ensure they eventually received care.

## DISCUSSION

Despite the limitations presented, the outcomes of this pilot highlight a workflow that enables students to meaningfully participate in patient care within the telehealth setting. Our efforts have since expanded to include 100 rising second-year medical students. With these additional pre-clinical participants, the telehealth elective has serviced over 2000 patients from the RWJMS outpatient practices. As patient volumes stabilize in the outpatient clinics, the focus of the elective will shift towards education through several modifications. For example, we plan to increase the number of students assigned to each attending physician and schedule explicit debrief sessions. This modification will maintain the practical aspects of the elective, increase uniformity among student experiences, and guarantee educational moments. To ensure student exposure to various patient populations, we will share our presented workflow with several specialty clinics that have already expressed interest in conducting a similar telehealth elective. We will continue to develop different and exciting modifications to bridge the return of learners to the clinical space.

After a successful four-week pilot of our outpatient telehealth elective, we started developing an inpatient telehealth initiative based on the Yale iCollaborative resources.^3^ This initiative will allow medical students to contribute to inpatient care remotely as practices and hospitals gradually reopen. The primary objective of this initiative is to introduce students to telehealth encounters with patients where appropriate without risk of exposure to SARS-CoV-2, while additional objectives require students to participate in rounds and practice case presentations, contribute to patient documentation, and act as patient liaisons after discharge to ensure patients successfully connect with the next phases of their care. Successful integration of this inpatient clinical curriculum will improve the capacity necessary for displaced students to engage with patients, albeit through a different medium.

These objectives ultimately reflect the evolving demand for non-traditional digital healthcare services in periods of both crises and normalcy. The American Medical Association (AMA) recently published a playbook of telehealth workflows in support of recent national policy changes to reduce unnecessary visits to healthcare settings and concomitant exposure risks to the Coronavirus.^4^ The American Association of Medical Colleges (AAMC) further recognizes the ability of telehealth to support longitudinal patient care models in their Core Entrustable Professional Activities for Entering Residency.^5^ Together, these AMA and AAMC documents establish a framework from which medical schools can build educational telehealth opportunities into their curriculums.

With the development and implementation of these new telehealth initiatives, we hope to expand the realm of possibility for clinical learning experiences in the virtual setting. These novel educational experiences have the potential to make our clinical curriculum more resilient during both this global pandemic and beyond.

## Data Availability

Survey materials and data spreadsheets are available upon request.

## Acknowledgements

We thank Daniel Levin and Valeriya Gershteyn for facilitating remote EMR access and workflow optimization, without whom such quick adoption would not have been possible. Thanks also to Drs. Amy Tyberg and Frank Sonnenberg for their support, and to Julie Lawson for her flexibility in helping integrate the workflow into our many practices.

## DISCLOSURES

### Ethical approval

This study was approved by the Rutgers Institutional Review Board (study ID number PRO2020001884).

### Funding/Support

none.

### Other disclosures

none.

### Disclaimers

none.

## Notes

### Competing Interest Statement

The authors have declared no competing interest.

### Funding Statement

No external funding was received for this study.

### Author Declarations

This study was approved by the Rutgers Institutional Review Board (PRO2020001884).

